# Tau PET visual reads find sources of tau not explained by typical Alzheimer disease pathophysiology

**DOI:** 10.1101/2022.12.20.22283743

**Authors:** Charles D. Chen, Maria Rosana Ponisio, Jordan A. Lang, Shaney Flores, Suzanne E. Schindler, Anne M. Fagan, John C. Morris, Tammie L.S. Benzinger

## Abstract

^18^F-flortaucipir-PET received FDA approval to visualize tauopathy in the brains of adult patients with cognitive impairment being evaluated for Alzheimer disease (AD). However, manufacturer’s guidelines for the visual interpretation of ^18^F-flortaucipir-PET differs greatly from how ^18^F-flortaucipir-PET has been measured in research settings using standardized uptake value ratios (SUVRs). How visual interpretation relates to ^18^F-flortaucipir-PET SUVR, CSF biomarkers, or longitudinal clinical assessment is not well understood. Here we compare these various diagnostic methods in participants enrolled in studies of aging and memory (n=189, of whom 23 were cognitively impaired). Visual interpretation had high agreement with SUVR (98.4%); discordant participants had hemorrhagic infarcts or atypical AD tauopathies. Visual interpretation had moderate agreement with CSF p-tau181 (86.1%). Two participants demonstrated ^18^F-flortaucipir uptake from meningiomas. Visual interpretation could not predict follow-up clinical assessment in 9.52% of cases. We conclude that close association between AD tauopathy and clinical onset in group-level studies does not always hold at the individual level, with discrepancies arising from atypical AD, vascular dementia, or frontotemporal dementia. A better understanding of relationships across imaging, CSF biomarkers, and clinical assessment is needed to provide appropriate diagnoses for these individuals.

## Introduction

The pathological hallmarks of Alzheimer disease (AD) are amyloid-β (Aβ) plaques and misfolded hyperphosphorylated tau neurofibrillary tangles (NFTs) [1,2]. *In vivo* evaluation of aggregated tau or associated pathophysiology in AD was first performed using immunoassays for cerebrospinal fluid (CSF) tau phosphorylated at position 181 (p-tau) [3]. Later, tau PET radiotracers were developed [4–6], along with methods for tau PET standardized uptake value ratio (SUVR) analyses [7,8]. The first generation of tau PET radiotracers includes the arylquinoline derivatives ^18^F-THK5317 and ^18^F-THK5351, the pyrido-indole derivative ^18^F-flortaucipir, and the phenyl/pyridinyl-butadienyl-benzothiazole/benzothiazolium derivative ^11^C-PBB3. Among these, ^18^F-flortaucipir (Tauvid™, Avid Radiopharmaceuticals) became the first to be approved by the United States Food and Drug Administration to estimate the density and distribution of aggregated tau NFTs in adult patients with cognitive impairment being evaluated for AD. Following the manufacturer’s guidelines for performing a visual interpretation of ^18^F-flortaucipir PET imaging involves identifying the presence or absence of confluent radiotracer uptake greater than 1.65 times the cerebellar uptake in either the posterolateral temporal, occipital, or parietal/precuneus regions. This method differs greatly from most research procedures for automated quantification of tau PET imaging data, such as taking the volume-weighted mean standardized uptake value ratio (SUVR) in a temporal meta-region of interest (ROI) and comparing that to a cohort-defined threshold [7,8]. These methodological differences may lead to disagreements between visual interpretation and SUVR quantification. In particular, the temporal meta-ROI used in SUVR quantification does not contain any of the occipital or parietal/precuneus structures used in visual interpretation, and includes several medial temporal lobe structures ignored in visual interpretation. Additionally, in the clinic ^18^F-flortaucipir PET imaging is only indicated for use in adult patients with cognitive impairment who are being evaluated for AD, whereas in a research setting ^18^F-flortaucipir PET imaging is performed regardless of cognitive status, calling into question whether ^18^F-flortaucipir PET imaging is a reliable measure of tauopathy during preclinical stages of AD. Tau pathophysiology can also be evaluated by measuring phosphorylated tau concentrations in the CSF, and several studies have provided additional evidence that tau PET is more strongly coupled to cognitive decline, whereas CSF p-tau181 is more tightly linked to preclinical AD [9–12]. Understanding where these three methods – tau PET visual interpretation, tau PET SUVR quantification, and CSF p-tau181 concentration – agree and differ may improve how we define AD tauopathy and AD clinical diagnoses.

## Materials and Methods

### Study participants

Participants selected for this study were enrolled in longitudinal observational studies of aging and memory at the Charles F. and Joanne Knight Alzheimer Disease Research Center (Knight ADRC, n=189, of whom 23 were cognitively impaired, Table 1). All participants met the inclusion criteria of having a tau PET usable for visual reads, and an Aβ PET, MRI, and clinical and cognitive evaluation, all within 18 months; the majority of participants (n=144) also underwent lumbar puncture within 18 months of their tau PET scan. The study was approved by the institutional review board, and all participants or their designees signed an informed consent form.

**TABLE 1.** Participant characteristics

|  |  | Cognitively normal | Cognitively impaired | Total |
| --- | --- | --- | --- | --- |
| **Number** |  | 166 | 23 | 189 |
| **Mean age in years (SD)** | | 68.9±8.34 | 75.7±7.36 | 69.8±8.51 |
| **Female (%)** |  | 93 (56.0) | 12 (52.2) | 105 (55.6) |
| **Race** | **White** | 147 | 23 | 170 |
|  | **Black or African American** | 18 | 0 | 18 |
|  | **Asian** | 1 | 0 | 1 |
| **Mean MMSE (SD)** |  | 29.2 (1.12) | 26.0 (3.66) | 28.8 (1.94) |
| **CDR<sup>®</sup>** | **0** | 166 | 0 | 166 |
|  | **0.5** | 0 | 16 | 16 |
|  | **1** | 0 | 6 | 6 |
|  | **2** | 0 | 1 | 1 |
| **Clinical diagnosis** | **Cognitively normal** | 166 | 0 | 166 |
|  | **Uncertain dementia** | 0 | 9 | 9 |
|  | **0.5 in memory only** | 0 | 1 | 1 |
|  | **AD dementia** | 0 | 13 | 13 |
| ***APOE* genotype** | **2/2** | 1 | 0 | 1 |
|  | **2/3** | 27 | 1 | 28 |
|  | **2/4** | 6 | 1 | 7 |
|  | **3/3** | 83 | 8 | 91 |
|  | **3/4** | 42 | 11 | 53 |
|  | **4/4** | 6 | 2 | 8 |
|  | **Unknown** | 1 | 0 | 1 |
| **Tau PET temporal meta-ROI SUVR** | **Mean±SD** | 1.15±0.106 [0.924, 1.882] | 1.44±0.364 [1.024, 2.43] | 1.18±0.185 [0.924, 2.43] |
|  | **Positive (%)** | 4 (2.41) | 13 (56.5) | 17 (8.99) |
| **Tau PET visual** | **Positive (%)** | 6 (3.61) | 14 (60.9) | 20 (10.6) |
| interpretation |  |  |  |  |
| Aβ PET (Centiloid) | Mean±SD | 19.9±34.4 | 74.3±45.6 | 26.5±40.0 |
|  | Positive (%) | 45 (27.1) | 19 (82.6) | 64 (33.9) |

Abbreviations: CDR<sup>®</sup>=Clinical Dementia Rating<sup>®</sup>, MMSE=Mini-Mental State Exam, SD=standard deviation.

### Clinical and cognitive assessment

Participants were assessed clinically and cognitively using the neuropsychological test battery from the Uniform Data Set (UDS) [13], which includes the Clinical Dementia Rating (CDR^®^) [14] and the Mini Mental State Examination (MMSE) [15]. The CDR assesses three domains of cognition (memory, orientation, judgment and problem solving) and three domains of function (community affairs, home and hobbies, personal care): scores from the six domains can either be summed to yield the CDR Sum of Boxes score, or passed to a lookup table to yield the CDR Global score.

### Tau PET acquisition

Participants were scanned on a Siemens Biograph 40 TruePoint (Siemens Healthineers). Participants received a single intravenous bolus injection (341±29.8 MBq) of ^18^F-flortaucipir (Tauvid™, Avid Radiopharmaceuticals). Emission data were collected 80-100 minutes post injection. List-mode data were reconstructed using ordered subset expectation maximization with three iterations and 21 subsets. A low-dose CT scan preceded PET acquisition for attenuation correction.

### Tau PET SUVR

Reconstructed PET images were processed using the PET Unified Pipeline (https://github.com/ysu001/PUP) and coregistered to corresponding MR images [16,17]. After segmenting MR images into regions of interest (ROIs) using FreeSurfer version 5.3 [18], regional SUVRs were defined from the reconstructed PET images using a cerebellar gray reference region. The temporal meta-ROI SUVR was defined as the volume-weighted mean SUVR of the amygdala, entorhinal, fusiform, parahippocampal, inferior temporal, and middle temporal ROIs [7,8].

### Tau PET visual interpretation

Two radiologists with training in nuclear medicine (J.A.L. and M.R.P.) followed the manufacturer’s guidelines for ^18^F-flortaucipir PET visual interpretation of participant scans using MIM Encore (MIM Software). Reconstructed PET images were coregistered with corresponding MR images. A ROI was drawn around the whole cerebellum in the axial plane that maximizes its cross-sectional area. A color scale with a rapid transition at 1.65 times the mean cerebellar counts was defined. The temporal lobe was divided into the anterolateral, anterior mesial, posterolateral, and posterior mesial temporal quadrants by placing the horizontal crosshair posterior to the brainstem nuclei, and the vertical crosshair at the widest portion of the temporal pole. An image was considered positive if it showed confluent activity above the rapid transition/cutoff in the cortical gray matter of the posterolateral temporal, occipital, or parietal/precuneus regions. An image was considered negative if it showed no activity above the cutoff in the cortical gray matter of the posterolateral temporal, occipital, or parietal/precuneus regions, or if it showed activity above the cutoff in the cortical gray matter restricted to the medial temporal, anterolateral temporal and frontal regions. Off-target binding, which may be seen in the choroid plexus, striatum, and brainstem nuclei, and small foci of nonconfluent activity, which may be seen throughout the cortical gray matter, were not used when determining tau positivity. Radiologists were blinded to all other information about each participant. In addition to following the manufacturer’s guidelines for ^18^F-flortaucipir PET visual interpretation, in this study, radiologists also reported whether radiotracer activity was symmetric across left and right hemispheres and whether there was off-target binding in the choroid plexus, striatum, brainstem nuclei, or bone/meninges. Notable findings (such as incidental meningiomas) were also reported.

### Aβ PET

Participants were scanned on either a Siemens Biograph 40 TruePoint, Biograph mMR, or Biograph Vision 600 (Siemens Healthineers). Participants received either a single intravenous bolus injection (539±159 MBq) of ^11^C-Pittsburgh compound B (PiB) or (369±22.4 MBq) of ^18^F-florbetapir (Amyvid™, Avid Radiopharmaceuticals). Emission data were either collected 30-60 minutes post injection (^11^C-PiB) or 50-70 minutes post injection (^18^F-florbetapir). Reconstructed PET images were formed and pre-processed in the same manner as tau PET. An Aβ PET SUVR was defined for each radiotracer [16,17] and standardized to the Centiloid scale [19,20].

### MR acquisition

Participants were scanned on either a Siemens Biograph mMR or Magnetom Vida (Siemens Healthineers). Across all scanners, T1-weighted head MR images were acquired using a magnetization prepared rapid gradient echo (MPRAGE) generalized autocalibrating partial parallel acquisition (GRAPPA) sequence using a repetition time=2300 ms, echo time=2.95 ms, flip angle=9°, at 1.1×1.1×1.2 mm^3^ voxel resolution.

### CSF

CSF was collected under standardized operating procedures. Participants underwent lumbar puncture in the morning following overnight fasting and 20-30 ml of CSF was collected in a 50 ml polypropylene tube via gravity drip using an atraumatic Sprotte 22-gauge spinal needle. CSF samples were kept on ice and centrifuged at low speed within two hours of collection, then transferred to another 50 ml tube to remove cells. CSF was aliquoted at 500 μl into polypropylene tubes and stored at -80°C. Concentrations of CSF p-tau181, Aβ42, and Aβ40 were measured by chemiluminescent enzyme immunoassay using a fully automated platform (LUMIPULSE G1200, Fujirebio) according to the manufacturer’s specifications.

### Statistical analyses

Cutoffs for binarizing tau PET, Aβ PET, CSF p-tau181, and CSF Aβ42/Aβ40 values were determined by fitting a two-component univariate Manly mixture model [21] in R software [22] to all relevant baseline PET SUVR or CSF measurements available in the Knight ADRC Data Freeze 17 and finding the decision boundary. Manly mixture modeling was used to account for possible severe skewness in the data that would be difficult to model using Gaussian mixture modeling, and to account for skewness that can vary from component to component, which would be impossible to model using log or Box-Cox transformations [21]. Cohen’s kappa (κ) was used to measure inter-rater reliability between the two radiologists’ tau PET visual interpretations, as well as between tau PET visual interpretation and tau PET SUVR quantification, and between tau PET visual interpretation and CSF p-tau181 concentration.

## Results

Overall, participants were on average (±standard deviation) 69.8±8.51 years old, most were cognitively normal with a global Clinical Dementia Rating (CDR^®^) [14] of 0 (n=166/189, 87.8%) and most did not carry the *APOE* ε4 allele (n=120/188, 63.8%). (Table 1). Cognitively normal participants had a mean tau PET temporal meta-ROI SUVR of 1.15±0.106 and a mean Centiloid of 19.9±34.4. Cognitively impaired participants (n=23/189, 12.2%) had a clinical diagnosis of either uncertain dementia (n=9), a CDR=0.5 in memory only (n=1), or AD dementia (n=13). They also had a mean tau PET temporal meta-ROI SUVR of 1.44±0.364 and a mean Centiloid of 74.3±45.6.

To binarize Aβ and tau biomarker data, Manly mixture modeling was used to determine the following cutoffs: tau PET temporal meta-ROI SUVR cutoff=1.32, Aβ PET (Centiloid) cutoff=21.6, CSF p-tau181 cutoff=58.1 pg/ml, and CSF Aβ42/Aβ40 cutoff=0.0737.

Of the 189 ^18^F-flortaucipir PET images, 20 (10.6%) were read as positive by both radiologists. Both radiologists also read 169 images as negative and thus agreed on the overall visual interpretation of each image in the current study (n=189/189, 100%, κ=1). Agreement between visual interpretation and SUVR quantification was high (n=186/189, 98.4%, κ=0.910) (Figure 1).

**FIGURE 1.**
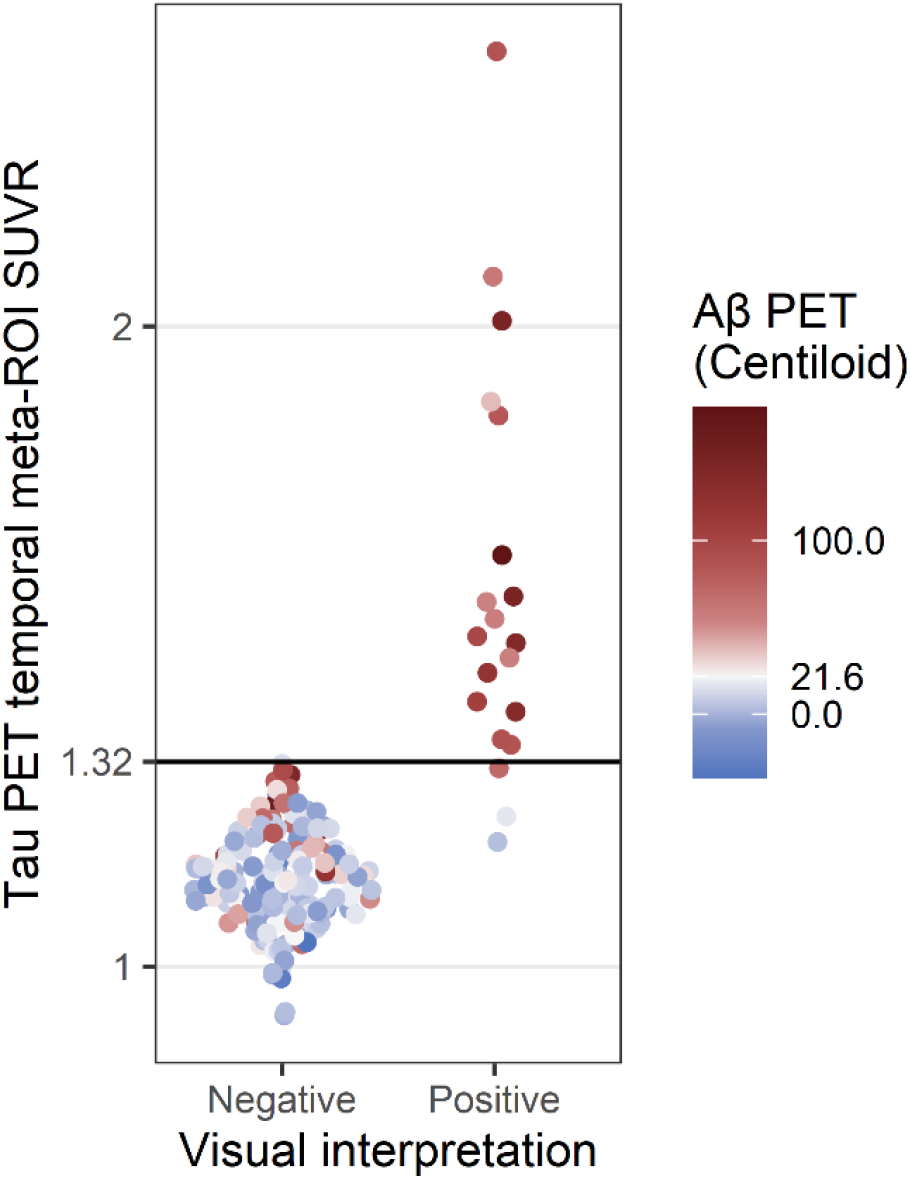
Comparison of tau PET visual interpretation with tau PET SUVR. Each PET study was assessed by visual interpretation using the manufacturer’s guidelines to determine positivity (x-axis) and by temporal meta-ROI SUVR analysis using a cutoff of SUVR=1.32 to determine positivity (y-axis). The color indicates the Aβ PET status for each case (positive Aβ PET, red; negative Aβ PET, blue; cutoff=21.6 Centiloids).

The three participants who had discordant results between visual interpretation and SUVR quantification all had tau-positive visual interpretations and tau-negative SUVRs. One participant (Figure 2a) demonstrated elevated ^18^F-flortaucipir uptake in the right precuneus and was Aβ PET, CSF Aβ42/Aβ40, and CSF p-tau181 negative (Table 2). Additional MR imaging revealed a hypointensity on T2*-weighted MRI that colocalized with the elevated right precuneus radiotracer uptake on ^18^F-flortaucipir PET (Figure 2b). The other two participants (Figures 2c and 2d) demonstrated lateralized occipital uptake, with greater uptake in either the left (Figure 2c) or right (Figure 2d), and were Aβ PET, CSF Aβ42/Aβ40, and CSF p-tau181 positive (Table 2). The participant with right occipital uptake also had posterolateral temporal and parietal/precuneus uptake.In terms of incidental findings, frontal meningiomas were identified in two participants. One participant had a meningioma in their left posterior frontal lobe (Figure 3a and 3b); the other participant had it in their left frontal lobe (Figure 3c and 3d). Both meningiomas had elevated levels of radiotracer uptake. The first participant also had elevated right posterolateral temporal uptake and tau-positive visual interpretation and SUVR and was Aβ PET positive (Table 3). The other participant had tau-negative visual interpretation and SUVR and was Aβ PET negative.

**TABLE 2.** AD biomarker status for cases with positive tau PET visual interpretation but negative tau PET SUVR analysis

| | Age | Sex | APOE | CDR<sup>®</sup> | Aβ PET<br>(Centiloid) | Tau PET<br>(SUVR) | CSF<br>Aβ42/Aβ40 | CSF p-tau181<br>(pg/ml) |
| --- | --- | --- | --- | --- | --- | --- | --- | --- |
| **Parietal/precuneus hemorrhagic infarct** | 80s | Male | 3/4 | 0 | 3.87 | 1.19 | 0.0975 | 21.6 |
| **Left occipital** | 70s | Female | 3/4 | 0 | 17.0→**50.0\*** | 1.23 | **0.0523** | **69.2** |
| **Right occipital** | 70s | Female | 3/3 | 0.5 | **72.1** | 1.31 | **0.0493** | **63.5** |

Numbers in bold denote positive biomarker status.
\*This participant had a Centiloid=17.0 (below cutoff) approximately one year before their tau PET visit, and a Centiloid=50.0 (above cutoff) approximately two years after their tau PET visit.

**TABLE 3.** AD biomarker status for cases with incidental meningioma

| | Age | Sex | APOE | CDR<sup>®</sup> | Aβ PET<br>(Centiloid) | Tau PET<br>(SUVR) | CSF<br>Aβ42/Aβ40 | CSF p-tau181<br>(pg/ml) |
| --- | --- | --- | --- | --- | --- | --- | --- | --- |
| **Left posterior frontal meningioma** | 70s | Female | 2/4 | 0 | **177** | **1.64** | **0.0481\*** | **49.9\*** |
| **Left frontal meningioma** | 70s | Male | 2/3 | 0 | 8.94 | 1.16 | 0.0848* | 30.3* |

Numbers in bold denote positive biomarker status.
\*CSF lumbar punctures were performed approximately 10 years prior to tau PET

**FIGURE 2.**
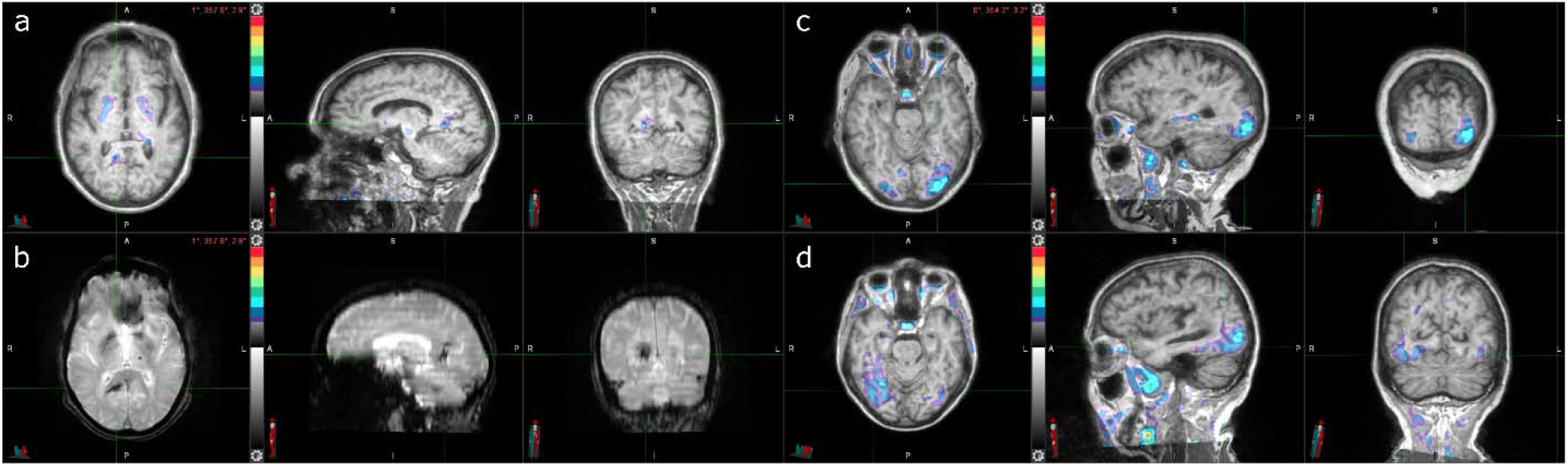
Three cases with tau-positive visual interpretations, but tau-negative SUVRs. (A) Tau PET coregistered with MRI of a man in his 80s with elevated right precuneus uptake. (B) Corresponding T2*-weighted MRI showing a hypointensity that colocalizes with the elevated right precuneus uptake from (A). (C) Tau PET coregistered with MRI of a woman in her 70s with elevated occipital lobe uptake, left greater than right. (D) Tau PET coregistered with MRI of a woman in her 70s with elevated posterolateral temporal, occipital, and parietal/precuneus lobe uptake, right greater than left.

**FIGURE 3:**
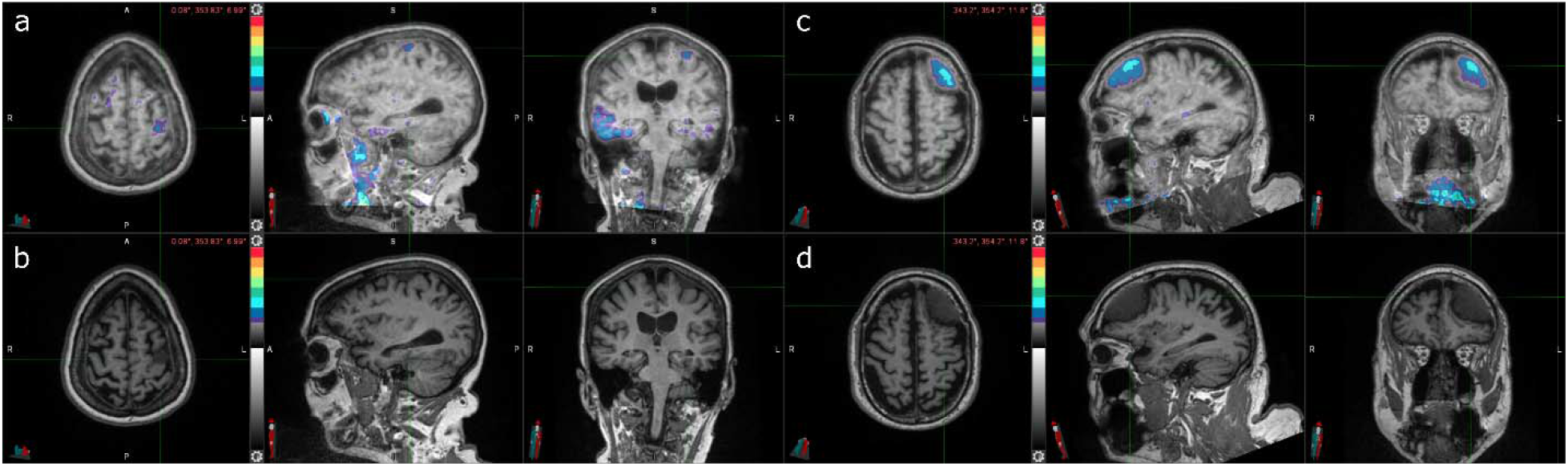
Two cases of incidental meningioma with tau PET uptake. (A) Tau PET coregistered with MRI of a woman in her 70s with a left frontal posterior meningioma with tau radiotracer uptake. (B) Corresponding stand-alone tau PET image. (C) Corresponding stand-alone MRI image. (D) Tau PET coregistered with MRI of a man in his 70s with a left frontal meningioma with tau radiotracer uptake. (E) Corresponding stand-alone tau PET image. (F) Corresponding stand-alone MRI image.

Agreement between visual interpretation and CSF p-tau181 was moderate (n=124/144, 86.1%, κ=0.526, Table 4). Two participants had tau-positive visual interpretations but were CSF p-tau181 negative (Figure 4a and 4b). One participant was previously identified as having a tau-positive visual interpretation but tau-negative SUVR (the same case as in Figure 2a and 2b). The other participant demonstrated posterolateral temporal uptake in both hemispheres and was Aβ PET and CSF Aβ42/Aβ40 positive. In addition, 18 participants had tau-negative visual interpretations but were CSF p-tau181 positive (Figure 4a and 4b). These cases were mostly Aβ PET positive (n=14/18, 77.8%) and/or CSF Aβ42/Aβ40 positive (n=17/18, 94.4%).

**TABLE 4.**
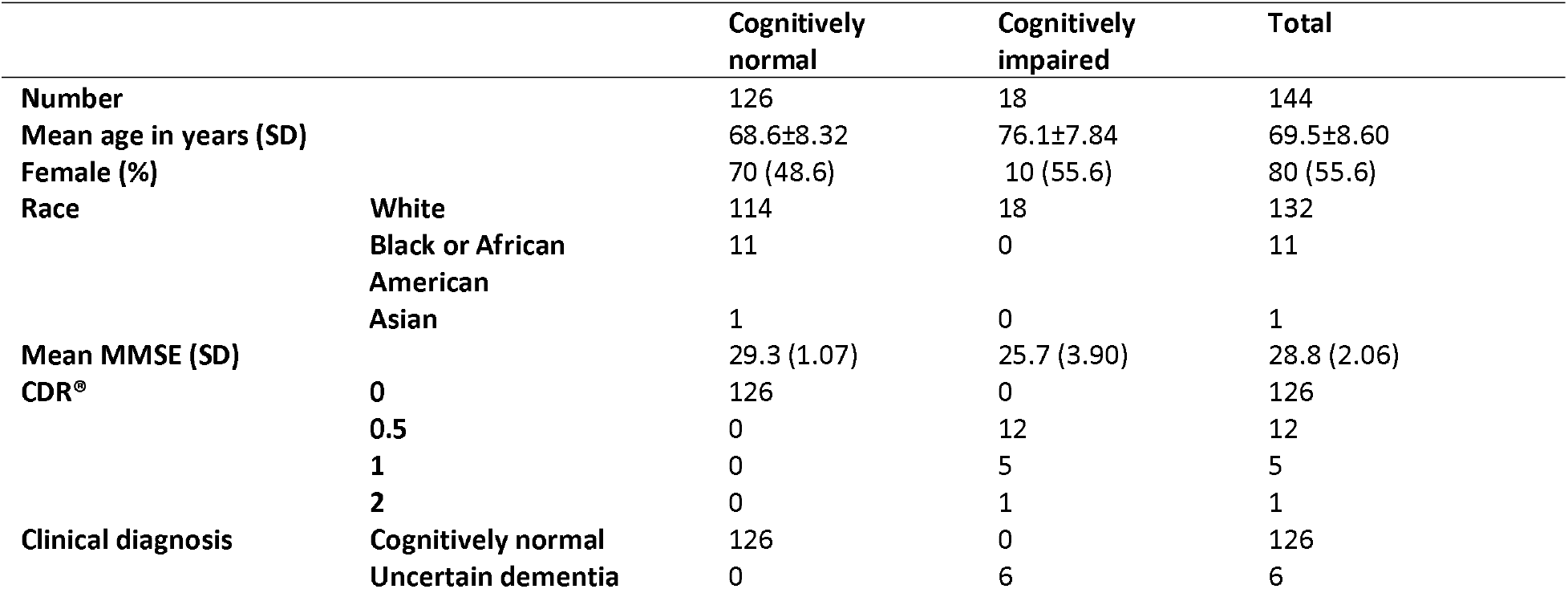
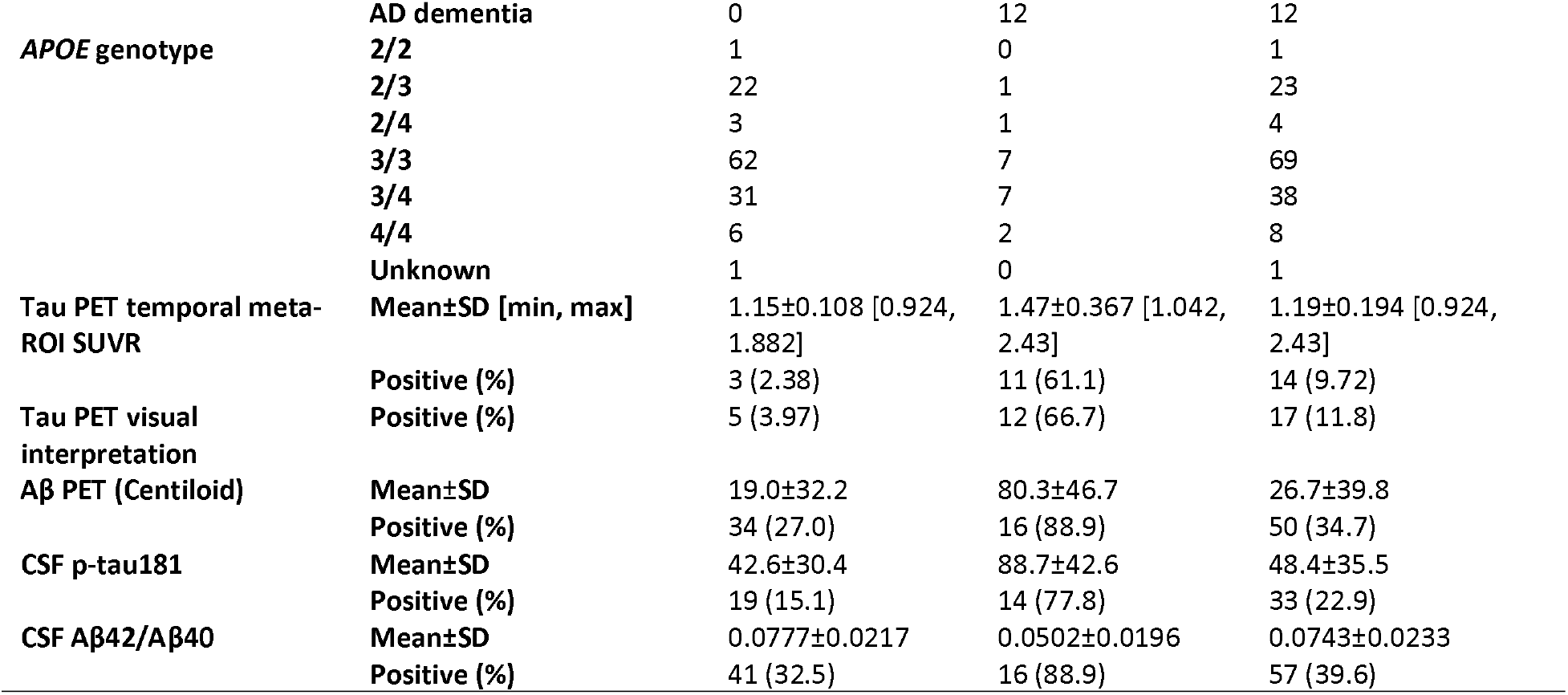
Participant characteristics for those who underwent lumbar puncture

**FIGURE 4:**
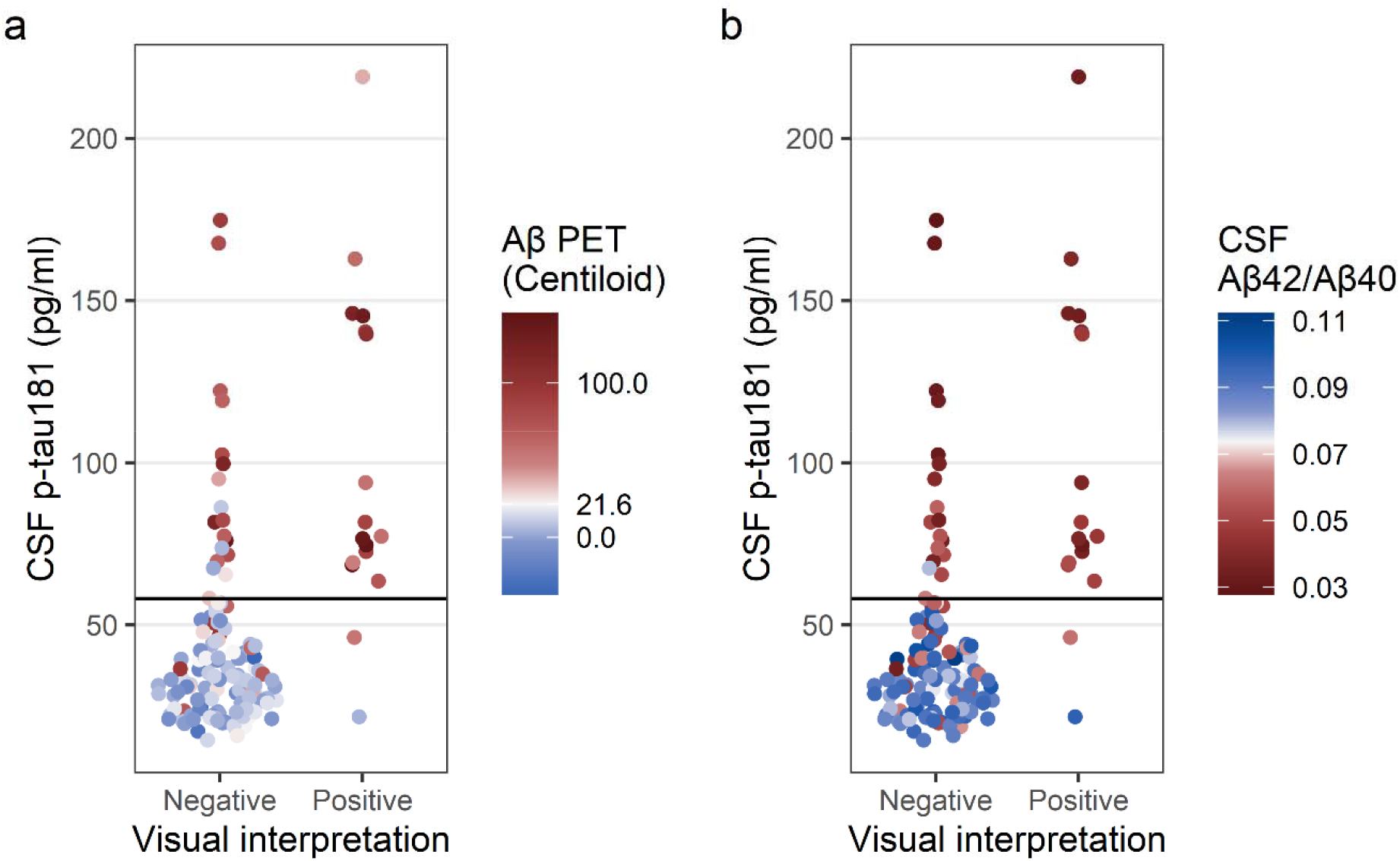
Comparison of tau PET visual interpretation with CSF p-tau181 concentration. (A and B) Each participant is plotted by visual interpretation (x-axis) and CSF p-tau181 concentration (y-axis); participants with p-tau181≥58.1 pg/ml were considered positive. In (A), the color indicates the Aβ PET status for each participant (positive Aβ PET, red; negative Aβ PET, blue; cutoff=21.6 Centiloid). In (B), the color indicates the CSF Aβ42/Aβ40 status for each participant (positive CSF Aβ42/Aβ40, red; negative CSF Aβ42/Aβ40, blue; cutoff=0.0737)

Six participants were assessed at baseline to be cognitively normal but tau-positive on visual interpretation (Table 5). One participant (Case 1) was previously mentioned to have PET radiotracer uptake colocalized to a parietal/precuneus hypointensity on T2*-weighted MRI and no other positive AD biomarkers (Figure 2 and Table 2). The remaining five participants were all Aβ PET positive. No participant reliably converted from cognitively normal to AD dementia. One participant (Case 2) did convert to AD dementia at their three-year follow up, but was reassessed to have a clinical diagnosis of uncertain dementia, more specifically, possible non-AD dementia of vascular origin at their five-year follow up. Another participant (Case 4) converted to AD dementia at their two-year follow up, but was reassessed to have frontotemporal dementia (FTD) at their four-year follow up.

**TABLE 5.** Cognitively normal participant follow up

|  | Age | Sex | APOE | Baseline tau PET temporal meta-ROI | Baseline tau PET visual interpretation | Asymmetry | Baseline Aβ PET (Centiloid) | Yearly follow-up clinical diagnosis |  |  |  |  |
| --- | --- | --- | --- | --- | --- | --- | --- | --- | --- | --- | --- | --- |
|  |  |  |  |  |  |  |  | 1 | 2 | 3 | 4 | 5 |
| Baseline cognitively normal and tau PET visual interpretation positive |  |  |  |  |  |  |  |  |  |  |  |  |
| 1* | 80s | Male | 3/4 | 1.19 | Positive | Right | 3.87 | CN | CN | CN | CN | CN |
| 2 | 70s | Male | 2/3 | 1.35 | Positive | Left | 86.6 |  | CN | AD | AD | UD |
| 3** | 70s | Female | 3/4 | 1.23 | Positive | Left | 17.0→50.0 | CN | CN | CN | CN | CN |
| 4 | 80s | Male | 3/3 | 1.88 | Positive | Left | 37.6 | UD | AD | AD | FTD |  |
| 5 | 80s | Male | 3/4 | 1.54 | Positive |  | 57.3 | CN |  |  |  |  |
| 6*** | 70s | Female | 2/4 | 1.64 | Positive | Right | 177 | CN | CN | CN |  |  |
| Baseline cognitively normal (and tau PET visual interpretation negative), but converts at follow up |  |  |  |  |  |  |  |  |  |  |  |  |
| 7 | 70s | Female | 3/4 | 1.22 | Negative |  | -2.35 | AD | CN | CN |  |  |
| 8 | 80s | Female | 3/4 | 1.21 | Negative |  | 120 | CN | AD | AD |  |  |
| 9 | 50s | Male | 3/3 | 0.93 | Negative |  | 3.38 |  |  | UD | AD |  |
| 10**** | 70s | Male | 2/3 | 1.16 | Negative |  | 8.94 | AD | CN | CN |  |  |

Numbers in bold denote positive biomarker status.
Abbreviations: AD=Alzheimer disease (dementia), CN=cognitively normal, FTD=frontotemporal dementia, UD=uncertain dementia. \*Same case as the “Parietal/precuneus hemorrhagic infarct” case in Table 2. \*\*Same case as the “Left occipital” case in Table 2. \*\*\*Same case as the “Left posterior frontal meningioma” case in Table 3. \*\*\*\*Same case as the “Left frontal meningioma” case in Table 3.

Four participants were assessed at baseline to be cognitively normal and tau-negative on visual interpretation, but would convert to AD dementia at follow up (Table 5). Two participants (Case 7 and Case 10) converted to AD dementia at their one-year follow ups, but were reassessed as cognitively normal at their two-year follow ups. The remaining two participants (Case 8 and Case 9) converted to AD dementia at their second- and fourth-year follow ups, respectively, but only Case 2 demonstrated Aβ PET positivity at baseline.

Twenty-three participants were assessed at baseline to have cognitive impairment (Table 6). Nine of these participants received a clinical assessment of uncertain dementia and two of the nine had a baseline tau-positive visual assessment. Both cases (Case 2 and Case 4) converted to AD dementia by their first- and second-year follow ups, respectively and were both Aβ PET positive. Nonetheless, three cases with a tau-negative visual interpretation at baseline (Case 5, Case 7, and Case 8) converted to AD dementia at their two-, two-, and three-year follow ups, respectively, although Case 5 was reassessed to be cognitively normal at their five-year follow up.

**TABLE 6.** Cognitively impaired participant follow up

|  | Age | Sex | APOE | Baseline tau PET<br>temporal meta-ROI | Baseline tau PET<br>visual interpretation | Asymmetry | Baseline Aβ PET<br>(Centiloid) | Yearly follow-up clinical diagnosis |  |  |  |  |
| --- | --- | --- | --- | --- | --- | --- | --- | --- | --- | --- | --- | --- |
|  |  |  |  |  |  |  |  | 1 | 2 | 3 | 4 | 5 |
| Baseline uncertain dementia |  |  |  |  |  |  |  |  |  |  |  |  |
| 1 | 60s | Male | 3/4 | 1.11 | Negative |  | 51.0 |  | UD | UD | CN | UD |
| 2 | 70s | Female | 3/4 | 1.86 | Positive |  | 82.7 | AD | AD | AD |  |  |
| 3 | 60s | Female | 3/3 | 1.14 | Negative | Right | 37.6 | CN |  | CN | CN | CN |
| 4* | 70s | Female | 3/3 | 1.31 | Positive | Right | 72.1 | UD | AD | AD | AD |  |
| 5 | 70s | Male | 3/4 | 1.10 | Negative | Right | 77.9 | CN | UD | AD | UD | CN |
| 6 | 70s | Female | 2/3 | 1.16 | Negative |  | -25.7 | UD |  |  |  |  |
| 7 | 80s | Male | 2/4 | 1.04 | Negative |  | 9.49 | CN | AD |  |  |  |
| 8 | 70s | Female | 4/4 | 1.29 | Negative |  | 80.1 |  | UD | AD | AD |  |
| 9 | 70s | Male | 3/4 | 1.02 | Negative | Right | 19.2 | CN | CN | CN | CN |  |
| Baseline 0.5 in memory only |  |  |  |  |  |  |  |  |  |  |  |  |
| 10 | 80s | Male | 3/3 | 1.07 | Negative |  | 12.2 | CN | CN |  |  |  |
| Baseline AD dementia |  |  |  |  |  |  |  |  |  |  |  |  |
| 11 | 70s | Female | 3/4 | 1.57 | Positive |  | 54.7 | AD | AD |  |  |  |
| 12 | 60s | Male | 3/3 | 2.43 | Positive |  | 85.7 |  |  |  |  |  |
| 13 | 70s | Female | 3/4 | 2.08 | Positive |  | 60.1 | AD | AD | AD | AD |  |
| 14 | 80s | Male | 3/3 | 1.46 | Positive |  | 121 | AD | AD |  |  |  |
| 15 | 80s | Male | 3/4 | 1.35 | Positive |  | 87.5 | AD | AD | AD | AD |  |
| 16 | 70s | Female | 4/4 | 1.58 | Positive | Left | 145 | AD | AD |  |  |  |
| 17 | 70s | Female | 3/4 | 1.52 | Positive |  | 97.2 | AD |  |  |  |  |
| 18 | 70s | Female | 3/4 | 1.18 | Negative |  | 62.8 | FTD |  |  |  |  |
| 19 | 70s | Male | 3/4 | 1.40 | Positive | Right | 134 | AD | AD | AD | AD |  |
| 20 | 80s | Male | 3/4 | 1.51 | Positive |  | 138 | AD | AD |  |  |  |
| 21 | 80s | Female | 3/3 | 1.41 | Positive | Right | 106 | CN | CN |  |  |  |
| 22 | 70s | Female | 3/4 | 1.48 | Positive | Left | 55.9 | AD | AD |  |  |  |
| 23 | 50s | Male | 3/3 | 2.01 | Positive |  | 145 |  |  |  |  |  |

Numbers in bold denote positive biomarker status.
Abbreviations: AD=Alzheimer disease (dementia), CN=cognitively normal, FTD=frontotemporal dementia, UD=uncertain dementia. \*Same case as the "Right occipital" case in Table 2.

Thirteen of the 23 participants with baseline cognitive impairment received a clinical assessment of AD dementia. All 13 participants were Aβ PET positive (Table 6). Twelve of these participants had tau-positive visual interpretation; the remaining participant (Case 18) was tau PET negative, but at their one-year follow up had their clinical assessment changed to frontotemporal dementia (FTD). Additionally, Case 21 was tau PET positive, but was reassessed to be cognitively normal at their one- and two-year follow ups.

## Conclusions

^18^F-flortaucipir PET visual interpretation was found to be consistent between readers in this study. However, three participants had discordant visual interpretations and SUVRs. In one participant, visual interpretation found elevated uptake in the right precuneus, which was missed by the temporal meta-ROI SUVR. However, when the tau PET images were coregistered with additional T2*-weighted MR imaging, the elevated radiotracer uptake was found to be colocalized with a T2* hypointensity, suggesting a hemorrhagic infarct to be the cause instead of AD tauopathy. All other AD biomarkers were negative for this participant, supporting this interpretation. Upon review of the additional T2*-weighted MR imaging, the readers also revised their interpretation of the image to be tau negative.

In another participant, visual interpretation found elevated uptake in the occipital lobe, with greater uptake in the left versus right hemisphere, which was also missed by the temporal meta-ROI SUVR. With all other AD biomarkers being positive for this participant, this participant likely has an occipital-predominant form of AD tau pathology [23].

Finally, in the third participant, visual interpretation found elevated uptake in the posterolateral temporal, occipital, and parietal/precuneus regions, with greater uptake in the right versus left hemisphere. The temporal meta-ROI SUVR was borderline negative, suggesting that, perhaps due to the lateralized uptake, the SUVR was artificially low for this case.

Among non-AD sources of ^18^F-flortaucipir uptake, the most studied is off-target binding in the choroid plexus, striatum, brainstem, and bone/meninges [24,25]. In this study, off-target binding did not mimic the appearance of the AD tau pattern when assessed by visual readers, nor did it cause any tau PET temporal meta-ROI SUVR to be falsely positive when compared to visual interpretation. However, we observed two other sources of off-target binding that were not mentioned in the manufacturer’s guidelines for ^18^F-flortaucipir PET visual interpretation and which can potentially confound tau PET interpretations: hemorrhagic infarcts and meningiomas. The hemorrhagic infarct case was the case previously described as having a tau-positive visual interpretation and a tau-negative SUVR quantification. The two meningioma cases demonstrated elevated levels of radiotracer uptake in the frontal lobe, which is immaterial when assessing tau PET positivity by visual interpretation, but meningiomas in the posterolateral temporal, occipital, or parietal/precuneus regions might plausibly interfere with visual interpretation and SUVR quantification.

When interpreting tau PET visual interpretation alongside clinical diagnosis after the study (both visual interpretation and clinical diagnosis were performed independently) a few relationships between the two kinds of AD diagnoses were remarkable. First, a baseline tau-positive visual interpretation in participants who were cognitively normal at baseline did not reliably predict conversion to AD dementia at follow up. If anything, tau PET positivity in cognitively normal participants was more likely to be either a sign of atypical AD, of related dementias (vascular dementia or FTD), or of resilience to AD dementia. Second, a baseline tau-negative visual interpretation in participants who were cognitively normal at baseline did not rule out conversion to AD dementia at follow up. Four cases were found to demonstrate conversion to AD dementia at follow up under these circumstances, although two of these were later reassessed to be cognitively normal. Third, baseline tau PET positivity in cognitively impaired participants did not guarantee a diagnosis of AD dementia at follow up: one participant was assessed to be cognitively normal at follow up even under these circumstances and another was reassessed to have FTD. Finally, baseline tau PET negativity in cognitively impaired participants cannot be used to rule out conversion to AD dementia at follow up: three such participants converted to AD dementia at their follow up visits, respectively, although one was reassessed to be cognitively normal at a later date.

A bias of the current study lies in the inclusion of cognitively normal participants. In a clinical setting, ^18^F-flortaucipir PET is indicated for use in patients with cognitive impairment. Two of the three cases discordant between visual interpretation and SUVR quantification in this study were from cognitively normal participants and would not warrant the use of ^18^F-flortaucipir visual interpretation in a clinical setting to begin with. Six of the 20 cases discordant between visual interpretation and CSF p-tau181 quantification were from cognitively normal participants and also would not warrant the use of ^18^F-flortaucipir visual interpretation in a clinical setting. Nonetheless, exploring tau positivity in cognitively normal participants in this study identified individuals who have atypical AD tau and clinical progression.

In conclusion, ^18^F-flortaucipir PET visual interpretation can identify atypical tauopathy that may be missed by SUVR quantification. However, while the manufacturer’s guidelines for ^18^F-flortaucipir PET visual interpretation address non-AD sources of uptake such as off-target binding, they do not address other non-AD sources of uptake such as hemorrhagic infarcts and meningiomas. Temporal meta-ROI SUVR was highly concordant with visual interpretation. However, SUVR analyses could not detect lateralized occipital-predominant AD tauopathy. CSF p-tau181 concentration was moderately concordant with visual interpretation and enabled detection of early changes in AD pathophysiology associated with tau hyperphosphorylation. However, these changes cannot be seen on PET. Finally, a positive visual interpretation did not make a follow up diagnosis of AD dementia inevitable, and a negative visual interpretation did not exclude the possibility of a follow up diagnosis of AD dementia. Additional work is needed to understand how multiple AD PET and CSF biomarkers might conceivably be used in tandem in a clinical setting alongside AD clinical evaluation in order to correctly diagnosis and treatment all individuals, not just those who demonstrate AD biomarker and clinical findings concordant with group-level trends.

## Data Availability

The datasets generated during and/or analyzed during the current study are available from the corresponding author on reasonable request.

## Acknowledgments

We thank the altruism of participants and their families and contributions of Knight ADRC support staff. Avid Radiopharmaceuticals provided technology transfer and precursor for ^18^F-flortaucipir and ^18^F-florbetapir. C.D.C. received support from the Knight ADRC T32 Fellowship (5T32AG058518-04) and the NSF GRFP Fellowship (DGE-1745038 and DGE-2139839). S.E.S. received support from NIA R01AG070941 and Barnes-Jewish Hospital Foundation. A.M.F. received support from various NIH grants (P30AG066444, P01AG003991, P01AG026276, U19AG032438). J.C.M. received support from various NIH grants (P30AG066444, P01AG003991, P01AG026276, U19AG032438).

## Author contributions

All authors contributed to the study conception and design. Data collection and analysis were performed by Charles D. Chen, Maria Rosana Ponisio, and Jordan A. Lang. The first draft of the manuscript was written by Charles D. Chen and all authors commented on previous versions of the manuscript. All authors read and approved the final manuscript.

## Disclosure and competing interests statement

C.D.C., J.A.L., and S.F. have no disclosures. M.R.P. received consulting fees from NFL Concussion Settlement Program; and received payment/honoraria from the University of Texas Medical School Galveston (Cooley Visiting Professor for the Department of Radiology). S.E.S. received support from NIA R01AG070941 and Barnes-Jewish Hospital Foundation; received payment/honoraria from the University of Wisconsin, University of Washington, University of Indiana, and St. Luke’s Hospital; is a board member of Greater Missouri Alzheimer’s Association; and received data on behalf of Washington University from C2N Diagnostics at no cost. A.M.F. received support from various NIH grants (P30AG066444, P01AG003991, P01AG026276, U19AG032438); received consulting fees from DiamiR and Siemens Healthcare Diagnostics Inc.; and participated on an advisory board at Roche Diagnostics, Genentech, and Diadem. J.C.M. received support from various NIH grants (P30AG066444, P01AG003991, P01AG026276, U19AG032438); received consulting fees from Barcelona Brain Research Center and TS Srinivasan Advisory Board; received payment or honoraria from Montefiore Grand Rounds and Tetra-Inst ADRC seminar series; and participated on an advisory board at Cure Alzheimer’s Fund. T.L.S.B. has investigator-initiated research funding from NIH, Alzheimer’s Association, Barnes-Jewish Hospital foundation, and Avid Radiopharmaceuticals (a wholly owned subsidiary of Eli Lilly); participates as a site investigator in clinical trials sponsored by Avid Radiopharmaceuticals, Eli Lilly, Biogen, Eisai, Janssen, and Roche; serves as an unpaid consultant to Eisai and Siemens; and is on the Speaker’s Bureau for Biogen.

## Notes

### Author Declarations

IRB of Washington University in St. Louis gave ethical approval for this work

